# Altered cerebellar white matter in sensory processing dysfunction is associated with impaired multisensory integration and attention

**DOI:** 10.1101/2020.11.11.20230219

**Authors:** Anisha Narayan, Mikaela A. Rowe, Eva M. Palacios, Jamie Wren-Jarvis, Ioanna Bourla, Molly Gerdes, Annie Brandes-Aitken, Shivani Desai, Elysa J. Marco, Pratik Mukherjee

**Affiliations:** Department of Radiology and Biomedical Imaging, University of California, San Francisco, CA, USA; Department of Medicine, Tulane University School of Medicine, New Orleans, LA, USA; Cortica Healthcare, San Rafael, CA, USA; Department of Neurology, University of California, San Francisco, CA, USA; Department of Bioengineering and Therapeutic Sciences, University of California, San Francisco, CA, USA

**Author notes:** **Correspondence:** Pratik Mukherjee, MD PhD, Center for Molecular and Functional Imaging, Department of Radiology and Biomedical Imaging, University of California, San Francisco, UCSF Box 0946, 185 Berry Street, Suite 350, San Francisco, CA 94107, USA. **Co-correspondence:** Elysa J. Marco, MD, Cortica Healthcare, 4000 Civic Center Dr, Ste 100, San Rafael, CA 94903, USA. These authors contributed equally to the manuscript (Anisha Narayan, Mikaela A. Rowe).

**Keywords:** Sensory processing, cerebellum, neurodevelopmental disorder, diffusion tensor imaging, connectivity, white matter, fractional anisotropy

## Abstract

Sensory processing dysfunction (SPD) is characterized by a behaviorally observed difference in the response to sensory information from the environment. While the cerebellum is involved in normal sensory processing, it has not yet been examined in SPD. Diffusion tensor imaging scans of children with SPD (n=42) and typically developing controls (TDC; n=39) were compared for fractional anisotropy (FA), mean diffusivity (MD), radial diffusivity (RD) and axial diffusivity (AD) across the following cerebellar tracts: the middle cerebellar peduncles (MCP), superior cerebellar peduncles (SCP), and cerebral peduncles (CP). Compared to TDC, children with SPD show reduced microstructural integrity of the SCP and MCP, which correlates with abnormal auditory behavior, multisensory integration, and attention, but not tactile behavior or direct measures of auditory discrimination. In contradistinction, decreased CP microstructural integrity in SPD correlates with abnormal tactile and auditory behavior and direct measures of auditory discrimination, but not multisensory integration or attention. Hence, altered cerebellar white matter organization is associated with complex sensory behavior and attention in SPD, which prompts further consideration of diagnostic measures and treatments to better serve affected individuals.

## 1 Introduction

Sensory processing dysfunction (SPD), also referred to as sensory integration disorder and sensory processing disorder, is a behaviorally described neurodevelopmental difference estimated to affect up to 16% of children in the general population, and 40-80% of children with other comorbid neurodevelopmental disorders, such as autism spectrum disorder (ASD) or attention deficit hyperactivity disorder (Ahn et al. 2004; Koziol, Budding, and Chidekel 2011). This increasingly recognized neurodevelopmental disorder affects the brain’s capacity to respond and adapt to a continuous flow of unimodal and/or multimodal sensory stimuli and significantly compromises learning and daily functioning (Miller 2009; Miller et al. 2007; Ryckman et al. 2017). Despite verbal and other intellectual performance abilities in the normal range, individuals with SPD can experience substantial difficulty with fine motor control, cognitive control, and behavioral regulation (Brandes-Aitken et al. 2018), leading to emotional and social difficulties including anxiety and negative self-image (McMahon et al. 2019; Miller 2009; Miller et al. 2007).

While SPD refers to an umbrella of phenotypic challenges (Miller et al. 2007), the most recognizable aspects of the condition are sensory over-responsivity, or hyperreactivity, and sensory under-responsivity, or hyporeactivity. The DSM-5 does not recognize SPD as a standalone disorder, though sensory hyper- and hypo-reactivity are listed as core symptoms of ASD (American Psychological Association 2013). Despite this overlap of SPD features and ASD, SPD also occurs in isolation in individuals who do not have the primary social communication challenges that form the ASD criteria (Ahn et al. 2004; Tavassoli et al. 2018). A 2018 study demonstrated that children who have SPD without ASD demonstrate higher empathy scores and less systematizing behavior than those with ASD (Tavassoli et al. 2018). In addition to behavioral features, SPD and ASD also differ with regard to certain neuroanatomical markers, including differences in white matter (WM) microstructural integrity of neural tracts that subserve socio-emotional processing (Chang et al. 2014). The findings from these previous studies provide evidence that atypical sensory reactivity is not simply a feature of the broader autism construct. However, our understanding of cerebellar differences in children with atypical neurodevelopment largely stems from the study of individuals with ASD.

Previous studies have demonstrated cerebellar differences between ASD and typically developing controls (TDC) at the cellular, structural, and functional level (Kern 2002; Koziol, Budding, and Chidekel 2011). Several diffusion tensor imaging (DTI) studies of ASD have shown alterations of WM microstructure in the pathways linking the cerebellum to the brainstem and cerebral hemispheres (Brito et al. 2009; Groen et al. 2011; Jeong et al. 2012; Shukla et al. 2010). A 2017 structural MRI study of individuals with ASD showed differences in morphological connectivity among neuroanatomical areas implicated in sensory processing, including sensory regions of the cerebral cortex, the amygdala, and the cerebellum, with altered connectivity between the cerebellum and cortical areas (Cardon, Hepburn, and Rojas 2017). Moreover, the cerebellum plays a role in multisensory processing, as its connectivity to the cerebral cortex is important for sensory integration across different modalities such as hearing, sight, touch and even smell (Cardon, Hepburn, and Rojas 2017; Sathyanesan et al. 2019; Zhao et al. 2018). While investigation of the structural and functional aspects of sensory processing has begun in children with isolated SPD and those with sensory over-responsivity and autism (Bar-Shalita et al. 2009; Brandes-Aitken et al. 2018; Marco et al. 2011; Molholm et al. 2020), there has so far been no exploration of the role of the cerebellum in children with isolated SPD. We posit that the cerebellum may contribute to acquisition, discrimination, modulation and integration of multisensory information for interpretation of the environment and generation of appropriate responses.

DTI is a neuroimaging method that measures the diffusion of water in the brain’s white matter. It allows us to visualize and quantify direction, uniformity, and orientation of WM tracts for the examination of structural connectivity, e.g., between cerebellum and sensory cortex areas in ASD (Mallott et al. 2019; Cardon, Hepburn, and Rojas 2017; Jeong et al. 2012). Thus, DTI measures can serve as a proxy for connectivity, especially in studying distributed neurological networks, and can reveal subtle microstructural anomalies undetectable by other noninvasive imaging methods (Payabvash et al. 2019). Commonly used DTI metrics include fractional anisotropy (FA), mean diffusivity (MD), radial diffusivity (RD), and axial diffusivity (AD; Alexander et al. 2007). Although prior studies have examined sensory processing using electroencephalography (EEG), magnetoencephalography (MEG), MRI, and functional MRI (fMRI), none to our knowledge have used DTI to study the cerebellum of children with SPD.

We know that children with SPD show reduced WM integrity, specifically in the posterior cerebral regions, correlating with atypical sensory processing behavior (Chang et al. 2014; Chang et al. 2016; Owen et al. 2013). However, there is an absence of research on hindbrain WM microstructure in SPD, specifically the brainstem and cerebellum. In this study, we use DTI to investigate the major output tracts of the cerebellum, the left and right superior cerebellar peduncles (SCP-L and SCP-R), as well as the major input tract to the cerebellum, the middle cerebellar peduncle (MCP), at its decussation. We also investigate two control WM regions in the brainstem, the left and right cerebral peduncles (CP-L and CP-R) at the midbrain, which contain cerebral fiber tracts that connect to subcortical structures in the lower brainstem and spinal cord. We hypothesize that the CP, SCP and MCP will all show the same increase of RD and, to a lesser extent, of MD, as well as a decrease of FA that we have previously demonstrated for cerebral WM tracts in SPD compared to typically developing children (Chang et al. 2016; Owen et al. 2013).

However, we postulate a dissociation in the relationship of WM microstructure in the cerebrum versus cerebellum with sensory processing and behavior. We hypothesize that the CP will most strongly correlate with unimodal sensory processing measures such as the Differential Screening Test for Processing (DSTP) and the Auditory and Tactile measures of the Sensory Profile, consistent with prior results from cerebral tracts in SPD (Chang et al. 2016; Owen et al. 2013). In contradistinction, because of the integral role of the cerebellum in multisensory integration (Cardon, Hepburn, and Rojas 2017; Sathyanesan et al. 2019; Zhao et al. 2018) and in attention (Ailion et al. 2020; Baumann et al. 2015; Buckner 2013), we hypothesize that the SCP and MCP will correlate more strongly than does the CP with measures of Multisensory Processing and Attentional Behavior from the Sensory Profile. Furthermore, we expect these correlations of cerebellar WM microstructure with sensory behavior to exist in the SPD cohort, but not in TDC who by definition do not exhibit atypical sensory processing or cognitive control.

## 2 Materials and Methods

### 2.1 Study design

Children 8 to 12 years of age were enrolled in this study from the University of California, San Francisco (UCSF) Sensory Neurodevelopment and Autism Program phenotype database (pediLAVA) and neuroimaging collection. The study design was approved by the UCSF Institutional Review Board and informed consents and assents were obtained from primary caregivers and study participants, respectively, in accordance with IRB policy. Consistent with previous studies in our lab, research designation of SPD was determined using the long-form Sensory Profile (SP; Dunn 1999), a parent report measure consisting of 125 questions about sensory behaviors with a 5-point Likert scale response. Totals in each sensory domain are then categorized into “Typical Performance,” “Probable Difference,” or “Definite Difference”. On the SP, higher scores are indicative of “Typical Performance”, while lower scores indicate increased sensory symptomatology, or a “Definite Difference”. SPD designation required a score in the “Definite Difference” range for at least one of the following sections of the SP: Auditory Processing, Visual Processing, Vestibular Processing, Tactile Processing, Multisensory Processing, or Oral Sensory Processing. To ensure all cases of SPD were recognized, a Short Sensory Profile (SSP) total score was calculated by adding the responses to the subset of 38 questions that make up the SSP. Those with a SSP total score in the “Definite Difference” range were also included in the SPD cohort. To be included in the TDC group, individuals could not meet any of the above criteria for a research diagnosis of SPD. The Social Communication Questionnaire (SCQ; Rutter, Bailey, and Lord 2003) parent report form was administered to all subjects to screen for ASD. Those scoring at or above the cutoff of 15 on the SCQ were then evaluated using the Autism Diagnostic Observation Schedule (ADOS; Lord et al. 1989). Individuals scoring above the ADOS diagnostic cutoff were excluded from analysis. Additional exclusion criteria included premature delivery (<37 weeks), brain malformation or injury, presence of a known genetic diagnosis (e.g. Fragile X syndrome), movement disorder, bipolar disorder, psychotic disorder, or hearing impairment.

### 2.2 Behavioral measures

In addition to informing research diagnostic criteria, the SP yields useful information about individual sensory domains and sensory-associated responses and behaviors. In order to explore correlations with DTI metrics, we obtained five domain-level subtotals from the SP: Auditory Processing, Visual Processing, Tactile Processing, Multisensory Processing, and Inattention/Distractibility.

Cognitive ability was assessed using the Wechsler Intelligence Scale for Children-Fourth Edition (WISC-IV; Wechsler 2003). To minimize potential confounding effects of intellectual disability and increase the likelihood of successfully scanning participants without sedation, inclusion was based on a Full-Scale IQ (FSIQ) ≥70. Additional measures of cognitive ability obtained from the WISC-IV include Verbal Comprehension Index (VCI), Perceptual Reasoning Index (PRI), Working Memory Index (WMI), and Processing Speed Index (PSI).

Finally, the Differential Screening Test for Processing (DSTP; Richard and Ferre 2006), a direct assessment used to identify problems with auditory processing at three levels—acoustic, acoustic linguistic, and linguistic—was administered to all subjects. The acoustic subtest assesses auditory processing skills not associated with language, including binaural integration of two different sets of numbers, recognition of patterns between musical tones, and discrimination of nonsense words against a background of white noise. The acoustic linguistic subtest assesses auditory processing associated with language and includes phonemic and phonic manipulation tasks. The linguistic subtest assesses understanding of semantics and pragmatics of language, such as knowledge of antonyms, prosodic interpretation, and the ability to guess a word based on descriptive clues. An overall DSTP Total Score was also calculated by totaling the three subtest scores. Because DSTP scores provide more objective measures of the specific aspects of auditory processing than the SP Auditory score, DSTP scores were also included in correlational analyses with DTI metrics.

### 2.3 Image acquisition protocol

Brain scans were performed using a 12-channel head coil on a 3-Tesla MRI scanner (Siemens Tim Trio; Erlangen, Germany). Whole-brain DTI scans were obtained using a diffusion-weighted echoplanar sequence with echo time = 8000 ms; repetition time = 109 ms, field of view = 220 mm; voxel size = 2.2 × 2.2 × 2.2 mm; 64 diffusion directions at b-value of 2000 s/mm^2^; and one brain volume at b-value of 0 s/mm^2^. T1-weighted images were acquired using 3-dimensional magnetization-prepared rapid acquisition gradient echo for anatomical registration (echo time = 2.98ms, repetition time = 2300ms, inversion time = 900 ms, flip angle = 9°).

### 2.4 DTI image processing

The FMRIB Software Library (FSL) version 5.0.8 (Oxford, UK) software was used for image processing. All steps of the image processing pipeline have been reported previously (Chang et al. 2016; Owen et al. 2013; Payabvash et al. 2019). First, the brain tissue was extracted using the Brain Extraction Tool (BET) from FSL. Then, the Diffusion Toolbox in FSL (DTIFIT) was used for preliminary data check, and to confirm that the principal eigenvectors (V1) were correctly oriented. Next, EDDY functions from FSL were consecutively applied to correct for susceptibility-induced distortion, eddy currents, and subject motion. After these corrections, the DTIFIT was applied on corrected diffusion scans to generate FA, MD, RD, and AD maps. The FA maps were first registered to the structural T1-weighted image by linear affine transformation; then the reverse matrix was used for registration of seed/target regions to original diffusion space. Five WM tracts were examined: John Hopkins University tracts of the SCP-L and SCP-R, the MCP at its decussation, and the CP-L and CP-R. The SCP, which contains WM fiber tracts that connect the cerebellum with the midbrain, and the MCP, which contains WM fiber tracts that originate in the pontine nuclei, were examined as they are the only outflow tract from the cerebellum and the largest inflow tract to the cerebellum, respectively. The CP was chosen as a control region, since it contains cerebral tracts and has not been previously studied in SPD. The inferior cerebellar peduncles were not analyzed due to their small size and location near the skull base.

### 2.5 Statistical analyses

All statistical analyses were conducted using SPSS statistical software. Group characteristics were compared using Student’s *t-*tests for continuous and Fisher’s exact-tests for nominal variables. Significance was determined at a 95% confidence interval. Two-tailed *t*-tests were run to determine group differences in AD, FA, MD, and RD values in the SCP-L and -R, MCP, and CP-L and -R tracts. Pearson correlations of FA, MD, and RD with the WISC-IV components (PRI, WMI, PSI, and FSIQ), DSTP components (acoustic, acoustic linguistic, and total score), and the SP subtests (auditory, visual, tactile, attention, and multisensory processing) in the five examined tracts were investigated by each group separately. Individual one-way ANOVAs were conducted to determine group interaction effects of significant correlations. Multiple comparison corrections were not performed because a prior hypothesis existed for each statistical test performed.

## 3 Results

### 3.1 Participant characteristics

A total of 42 children with SPD (13 female), and 39 TDC (8 female) were included in our analyses. *Table 1* provides their demographic and psychometric characteristics. Statistical analyses showed no significant differences between the SPD and TDC cohorts with regard to gender, age, or general cognitive performance. The cohorts did show significantly different PSI scores from the WISC-IV (*p = 0*.*02*).

**Table 1.** Subject characteristics. The first two columns provide group-level means with standard deviations in parentheses for age, verbal comprehension index (VCI), perceptual reasoning index (PRI), working memory index (WMI), processing speed index (PSI), and full scale IQ (FSIQ). For gender, the values listed in the first two columns are number of girls per group, with percentage of girls in parentheses. The third column lists p-values from group comparisons, with significant values at p<0.05 bolded.

|  | SPD (n=42) | TDC (n=39) | P-value |
| --- | --- | --- | --- |
| Age (years) | 9.85 (1.33) | 10.20 (1.08) | 0.207 |
| Gender (girl) | 13/42 (31%) | 8/39 (21%) | 0.320 |
| VCI | 117.19 (13.99) | 121.95 (12.57) | 0.118 |
| PRI | 112.17 (14.33) | 113.47 (12.63) | 0.670 |
| WMI | 108.05 (14.15) | 110.00 (10.77) | 0.493 |
| PSI | 93.79 (11.99) | 100.68 (13.39) | **0.020** |
| FSIQ | 111.88 (12.59) | 116.21 (10.24) | 0.097 |

### 3.2 DTI group differences between SPD and TDC

Using a threshold *p*-value of 0.05, analyses shown in *Figure 1* revealed significant differences between SPD and TDC groups in the SCP-R for MD, RD, and FA. There were no significant differences in the SCP-L; however, strong trends in the hypothesized direction were observed. The MCP showed significant group differences for MD and RD. The CP-R showed significant differences for FA and RD. Although not significant, trends in the hypothesized direction were seen for the CP-L. There were no significant group differences in AD in any of the white matter tracts examined. As expected, the SPD group showed lower FA and higher MD and RD values compared to the TDC group in all tracts with significant group differences in DTI metrics.

**Figure 1.**
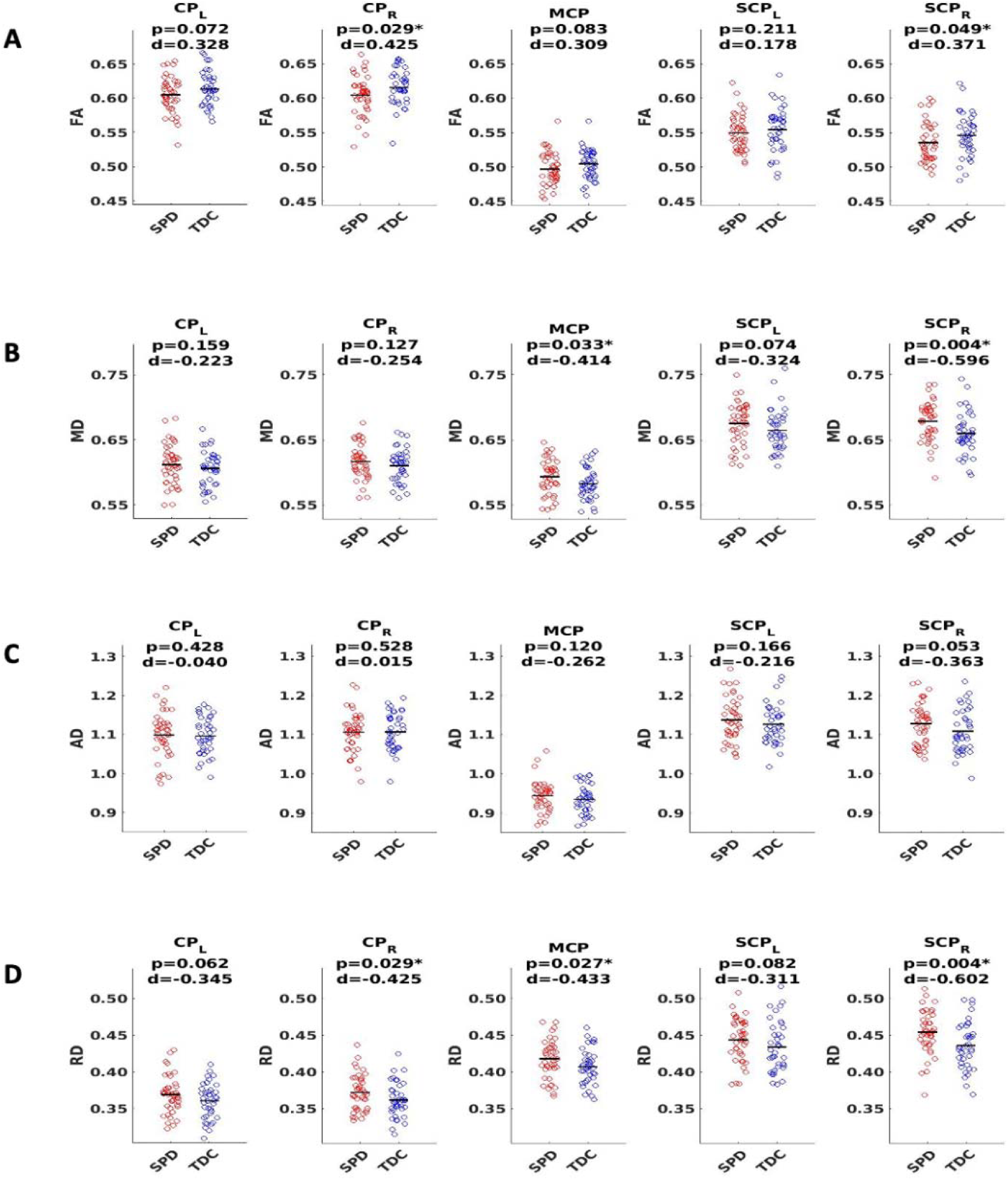
DTI group comparison of typically developing controls (TDC, blue) to sensory processing dysfunction subjects (SPD, red) using fractional anisotropy (FA), mean diffusivity (MD), axial diffusivity (AD) and radial diffusivity (RD). Tracts include the cerebral peduncles (CP, right and left), middle cerebellar peduncle (MCP), and superior cerebellar peduncles (SCP, right and left). (A) FA shows significant differences for CP-R and SCP-R, with lower FA in SPD subjects. (B) MD plots show significant differences for MCP and SCP-R, with higher MD in SPD subjects. AD plots show no significant differences for any of the five white matter tracts examined. (D) RD plots show significant differences for CP-R, MCP, and SCP-R with higher RD in SPD subjects. Statistical significance at p<0.05 is indicated by an asterisk.

### 3.3 DTI correlations with direct assessment of sensory measures

On the DSTP, a direct assessment of auditory discrimination, acoustic, acoustic linguistic, and DSTP total scores were significantly correlated with FA and RD in the CP-L and CP-R tracts in the SPD group but not the TDC group, consistent with our hypotheses (*Figure 2*). As expected, lower FA and higher RD were both associated with poorer auditory discrimination performance. However, no significant correlations were found between any of the DTI metrics examined and the DSTP linguistic score in either the SPD or TDC groups.

**Figure 2.**
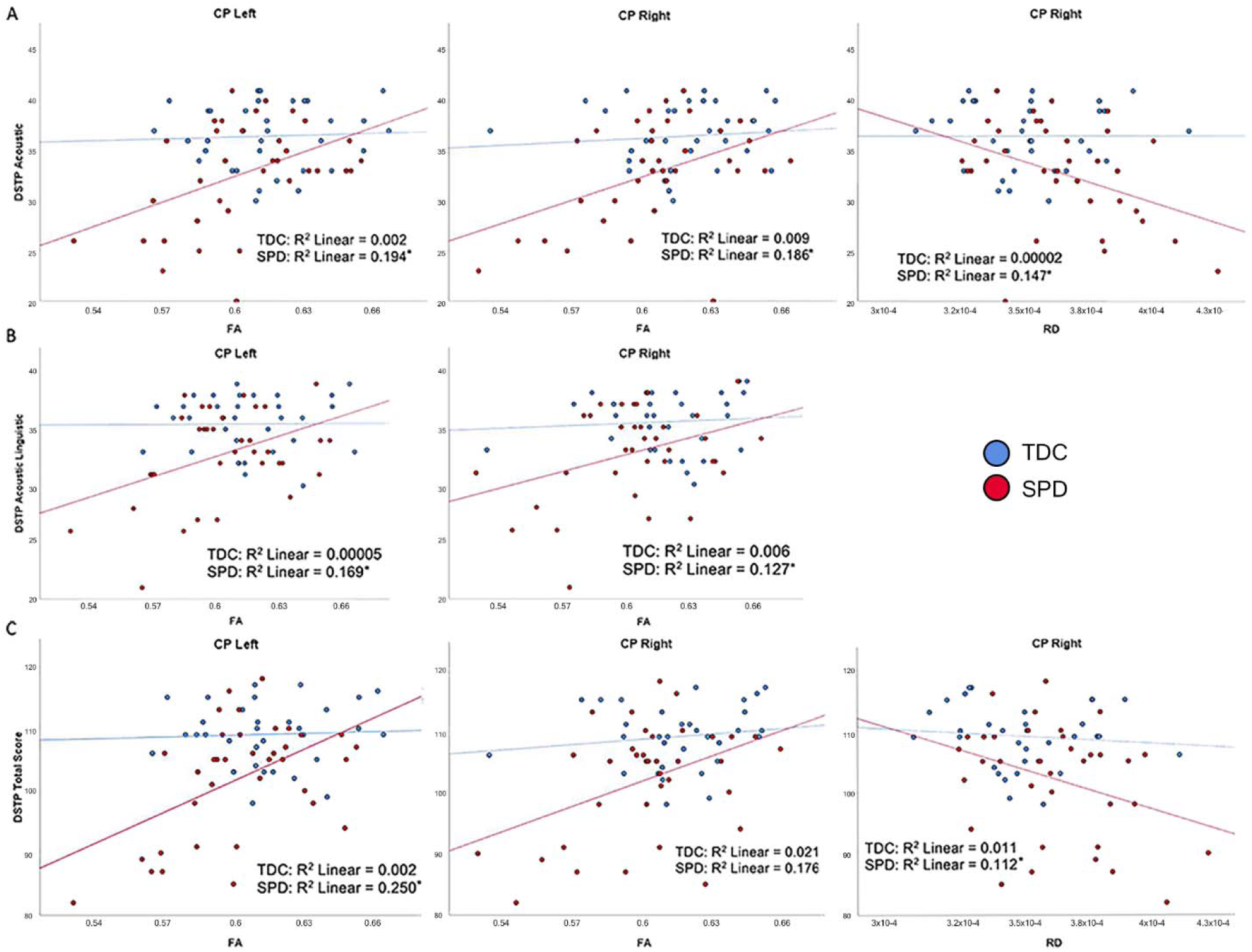
Correlation of TDC (blue) and SPD subjects (red) with Differential Screening Test for Processing (DSTP) scores. Statistical significance at p<0.05 is denoted with an asterisk next to the R2 value. (A) DSTP acoustic subtest score is correlated with FA in CP-L, FA in CP-R, and RD in CP-R for SPD but not TDC subjects. (B) DSTP acoustic linguistic subtest score is correlated with FA in CP-L and FA in CP-R for SPD but not TDC subjects. (C) DSTP total score is correlated with FA in CP-L, FA in CP-R, and RD in CP-R for SPD but not TDC subjects.

### 3.4 DTI correlations with parental assessment of unimodal sensory processing

The SP Auditory Processing subtotal significantly correlated with RD and MD values of all five tracts examined, which include both cerebellar and cerebral peduncles, in the SPD group (*Figure 3*). These results were consistent with the hypothesized relationship between higher diffusivities and more impaired sensory processing. Few significant correlations were observed in the TDC group, including RD in the MCP, where the correlation was reversed compared to the SPD group (*Figure 3*). Correlations that were significant for the TDC but not the SPD group included MD and RD in the CP-R and FA in the MCP. These correlations showed a similar pattern to RD in the MCP, where higher diffusivity correlated with less impaired sensory processing for TDC, the opposite relationship expected in the SPD group. This disparity in the correlation of DTI metrics with the SP Auditory Processing measure between the SPD and TDC groups is shown to be statistically significant as a group by tract interaction effect (*Table 2)*.

**Table 2.** Group, tract, and interaction effects of Sensory Profile scores and DTI metrics. P-values from one-way ANOVAs examining main effects of group, main effects of tract, and interaction effects for inattention, auditory, tactile, and multisensory processing subtotals from the SP. Significant values at p<0.05 are in bold.

| Metric | Tract | Inattention |  |  | Auditory Processing |  |  | Tactile Processing |  |  | Multisensory Processing |  |  |
| --- | --- | --- | --- | --- | --- | --- | --- | --- | --- | --- | --- | --- | --- |
|  |  | *Effect of cohort* | *Effect of tract* | *Interaction effect* | *Effect of cohort* | *Effect of tract* | *Interaction effect* | *Effect of cohort* | *Effect of tract* | *Interaction effect* | *Effect of cohort* | *Effect of tract* | *Interaction effect* |
| FA | CP-L | 0.055 | 0.922 | 0.152 | **0.025** | 0.993 | 0.080 | **0.003** | 0.102 | **0.010** | 0.208 | 0.659 | 0.383 |
|  | CP-R | **0.014** | 0.788 | 0.051 | **0.016** | 0.617 | 0.058 | **0.020** | 0.358 | 0.059 | 0.209 | 0.680 | 0.393 |
|  | MCP | **0.001** | 0.480 | **0.004** | **0.002** | 0.618 | **0.008** | 0.129 | 0.675 | 0.296 | 0.075 | 0.881 | 0.171 |
|  | SCP-L | **0.002** | 0.169 | **0.010** | 0.074 | 0.994 | 0.245 | 0.127 | 0.353 | 0.329 | **0.005** | 0.095 | **0.019** |
|  | SCP-R | **0.003** | 0.453 | **0.021** | 0.070 | 0.800 | 0.248 | **0.024** | 0.130 | 0.091 | **0.008** | 0.229 | **0.029** |
| MD | CP-L | 0.244 | 0.769 | 0.086 | **0.016** | 0.612 | **0.003** | 0.822 | 0.281 | 0.469 | 0.407 | 0.885 | 0.208 |
|  | CP-R | 0.157 | 0.794 | 0.055 | **0.017** | 0.563 | **0.004** | 0.708 | 0.656 | 0.941 | 0.664 | 0.596 | 0.409 |
|  | MCP | 0.204 | 0.286 | 0.076 | **0.049** | 0.785 | **0.013** | 0.769 | 0.969 | 0.870 | 0.408 | 0.450 | 0.224 |
|  | SCP-L | 0.095 | 0.070 | **0.025** | **0.040** | 0.359 | **0.009** | 0.765 | 0.366 | 0.420 | 0.288 | 0.201 | 0.134 |
|  | SCP-R | 0.321 | 0.167 | 0.124 | 0.354 | 0.639 | 0.132 | 0.644 | 0.265 | 0.340 | 0.528 | 0.198 | 0.293 |
| RD | CP-L | 0.209 | 0.752 | **0.039** | **0.025** | 0.566 | **0.002** | 0.169 | 0.077 | **0.035** | 0.399 | 0.969 | 0.141 |
|  | CP-R | 0.073 | 0.800 | **0.010** | **0.027** | 0.517 | **0.003** | 0.521 | 0.354 | 0.182 | 0.540 | 0.620 | 0.220 |
|  | MCP | 0.061 | 0.255 | **0.010** | **0.028** | 0.992 | **0.004** | 0.951 | 0.876 | 0.562 | 0.371 | 0.624 | 0.149 |
|  | SCP-L | **0.024** | **0.028** | **0.002** | 0.124 | 0.371 | **0.016** | 0.780 | 0.259 | 0.297 | 0.066 | **0.048** | **0.013** |
|  | SCP-R | 0.113 | 0.153 | **0.017** | 0.449 | 0.753 | 0.110 | 0.379 | 0.100 | 0.111 | 0.156 | 0.092 | **0.042** |

**Figure 3.**
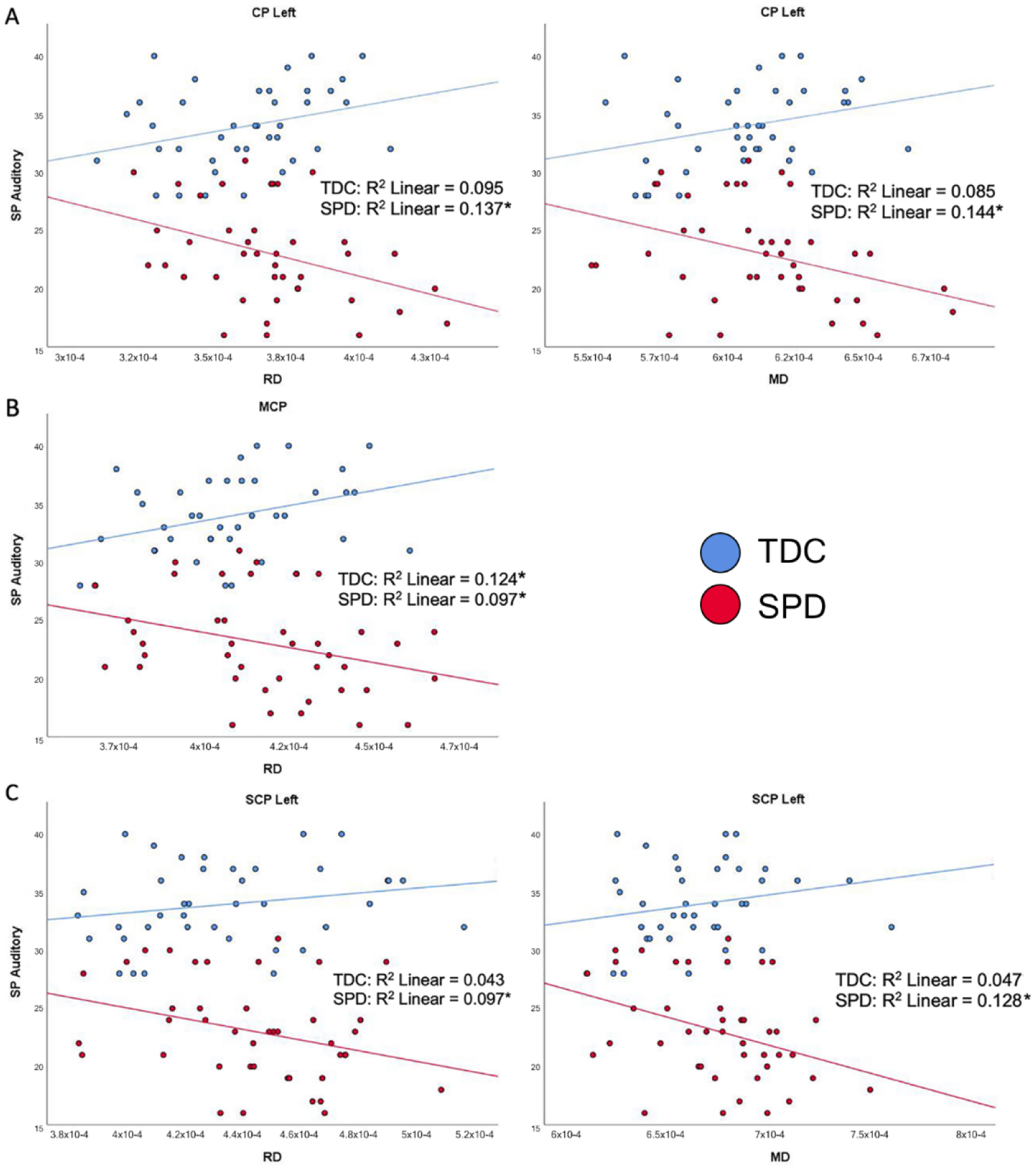
Correlation of TDC (blue) and SPD subjects (red) with the Sensory Profile (SP) auditory processing score. Statistical significance at p<0.05 is denoted with an asterisk next to the R2 value. (A) SP auditory processing score is correlated with RD in CP-L and MD in CP-L for SPD but not TDC subjects. (B) SP auditory processing score is correlated with RD in MCP for both SPD and TDC subjects. (C) SP auditory processing score is correlated with RD in SCP-L and MD in SCP-L in SPD but not TDC subjects.

In the SPD group, the SP Tactile Processing subtotal was significantly correlated with RD and FA of the CP-L (*Figure 4*), but none of the cerebellar tracts. No significant correlations were found in the TDC group. Therefore, significant group by tract interaction effects in tactile processing were identified for RD and FA of the CP-L (*Table 2*).

**Figure 4.**
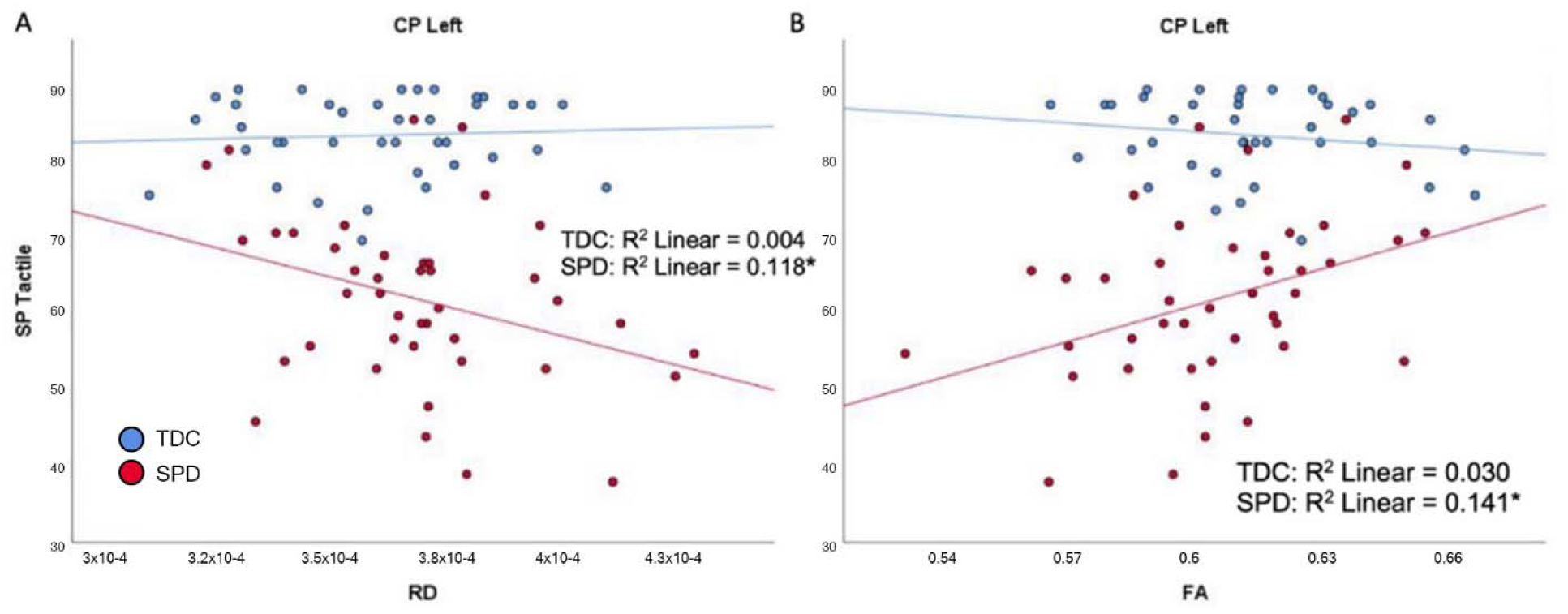
Correlation of TDC (blue) and SPD subjects (red) with SP tactile processing score. Statistical significance at p<0.05 is denoted with an asterisk next to the R2 value. (A) SP tactile processing score is correlated with RD in CP-L for SPD but not TDC subjects. (B) SP tactile processing score is correlated with FA in CP-L for SPD but not TDC subjects.

### 3.5 DTI correlations with parental assessment of multisensory integration and attention

For the SPD group, the SP Multisensory Processing subtotal was significantly correlated with RD and FA in the SCP-L and SCP-R (*Figure 5*). No significant correlations were found for the TDC group; hence, significant group by tract interaction effects were found for RD and FA of the SCP-L and SCP-R (*Table 2*). The SP Inattention subtotal was significantly correlated with RD, MD, and FA of the SCP-L and -R and the MCP in the SPD cohort, but not the TDC cohort (*Figure 6*). Significant group by tract interaction effects were therefore identified for RD and FA of all the cerebellar tracts (*Table 2*). Neither the SP Multisensory Processing nor the SP Inattention subtests showed any significant correlation with DTI metrics of the left or right cerebral peduncles for the SPD group, though the TDC group showed a significant positive correlation between RD in the CP-R and Inattention score. Once again, the relationship seen in the TDC group is opposite what we would expect to see in the SPD group.

**Figure 5.**
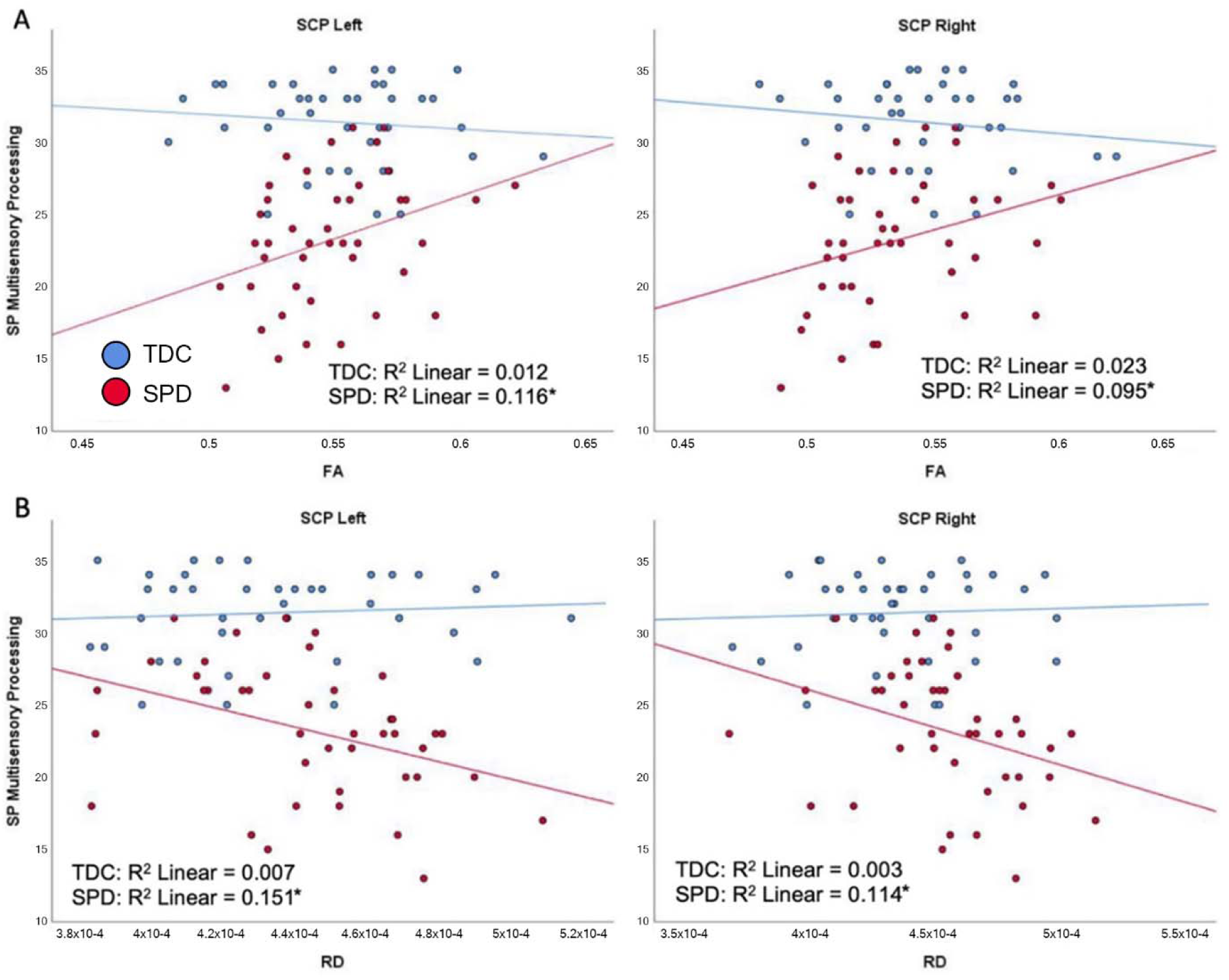
Correlation of TDC (blue) and SPD subjects (red) with SP multisensory processing score. Statistical significance at p<0.05 is denoted with an asterisk next to the R2 value. (A) SP multisensory processing score is correlated with FA in SCP-L and FA in SCP-R for SPD but not TDC subjects. (B) SP multisensory processing score is correlated with RD in SCP-L and RD in SCP-R for SPD but not TDC subjects.

**Figure 6.**
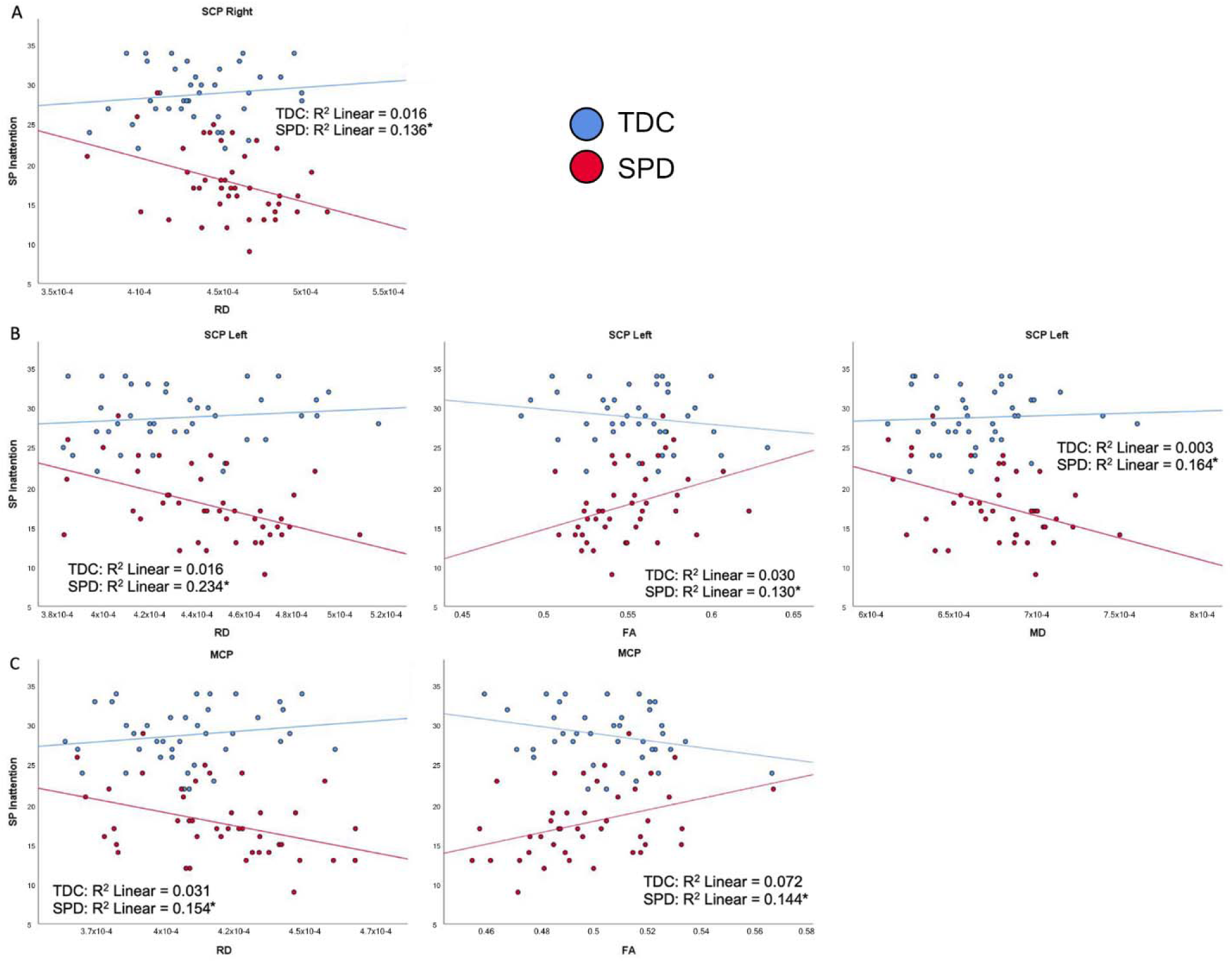
Correlation of TDC (blue) and SPD subjects (red) with SP inattention score. Statistical significance is denoted with an asterisk next to the R^2^ value. (**A**) Sensory Profile (SP) inattention score is correlated with RD in SCP-R for SPD but not TDC subjects. (**B**) SP inattention score is correlated with RD in SCP-L, FA in SCP-L, and MD in SCP-L for SPD but not TDC subjects. (**C**) SP inattention score is correlated with RD in MCP and FA in MCP for SPD but not TDC subjects.

## 4 Discussion

### 4.1 Alterations of brainstem and cerebellar white matter microstructure in SPD

Our results demonstrate that children with SPD have altered brainstem and cerebellar WM microstructural integrity relative to TDC. As hypothesized based on our previous studies of the cerebral hemispheres (Chang et al. 2016; Owen et al. 2013; Payabvash et al. 2019), these microstructural changes of the cerebral and cerebellar peduncles consist of reduced FA, increased MD, and increased RD. Consistent with these prior reports, RD typically showed the largest effect sizes, followed by FA and MD, whereas AD had the weakest effects. This supports the underlying biological principle that barriers to microscopic water diffusion radial to the orientation of axonal fiber bundles are not as well developed in SPD as they are for TDC. While recent studies have shown altered cerebellar connectivity in individuals with ASD (Cardon, Hepburn, and Rojas 2017; Oldehinkel et al. 2019), none have explored this relationship in children with SPD who do not have additional social-communication deficits meeting criteria for ASD. By examining an SPD cohort, we are able to investigate sensory processing dysfunction more directly, without the confounding effect of the additional brain-based cognitive and affective challenges. The fact that significant differences in cerebellar white matter microstructure exist between the two groups when isolating sensory dysfunction adds to increasing evidence that the cerebellum plays an important role in sensory processing and attention. Furthermore, evidence from a mouse model of white matter injury of prematurity shows a loss of GABA_A_ receptor–mediated synaptic input to cerebellar NG2 cells, resulting in extensive proliferation of these cells and delayed oligodendrocyte maturation, eventually leading to dysmyelination (Zonouzi et al. 2015). Prematurely born children demonstrate elevated rates of SPD (Wickremasinghe et al. 2013) and are known to have WM diffusivity changes on DTI that are characteristic of diminished myelination (i.e., elevated RD and MD as well as reduced FA) similar to those we demonstrate herein for the cerebellar and cerebral peduncles in term-born children with SPD. A new study of neurotypical children has shown a strong correlation between FA in the right and left SCP and quantitative T1 values, a more specific measure of myelination than DTI metrics, providing further support that our findings are at least in part due to myelination differences between groups (Bruckert et al. 2020). Thus, abnormal excitatory-inhibitory balance during cerebellar development may represent a possible mechanism for producing these microstructural WM alterations that are ultimately expressed as the SPD phenotype.

### 4.2 Cerebral versus cerebellar peduncle microstructure and unimodal sensory processing

Also consistent with our earlier findings about white matter of the cerebral hemispheres in SPD (Chang et al. 2016), WM microstructure of the left and right CPs in the SPD children was significantly correlated with all but the linguistic subtest scores of the DSTP, which is a direct assessment of auditory processing. In contradistinction, the SCPs and MCP in the SPD group were not associated with any of the DSTP scores, indicating that cerebellar white matter is not as directly involved in impaired auditory discrimination as is cerebral white matter.

Similar to the DSTP measures, the SP Tactile subtotal was associated with cerebral peduncle microstructure but not cerebellar peduncle microstructure. However, the SP Auditory subtotal was significantly correlated with WM microstructure in *both* cerebellar and cerebral peduncles. One possible explanation for this discrepancy is that the SP combines all aspects of auditory processing, including modulation, discrimination, and sensory-based motor issues, into a single auditory processing score, while the DSTP examines auditory discrimination specifically. However, Parsons et al. (2009) showed a relationship between impaired cerebellar functioning and poor performance on a pitch discrimination task, which does support the idea that the cerebellum may be important in some aspects of auditory discrimination. Another explanation is that certain questions in the SP involve both auditory and attention elements, such as “Is distracted or has trouble functioning if there is a lot of noise around”, which makes it difficult to assess auditory processing and attention separately using only the SP. Finally, these results may reflect the fact that the SP is a subjective parent report measure involving a behavioral questionnaire and the DSTP is a more objective, direct assessment. Previous research has shown that direct assessment of sensory processing is more strongly correlated with measures of cerebral WM microstructure than are parent report measures, which could potentially be due to parental bias (Chang et al. 2016).

RD showed the most consistent relationships with DSTP as well as SP Auditory and Tactile measures in the SPD cohort, followed by FA and MD. As expected, AD did not show any significant associations with any of the sensory measures in either the cerebral or cerebellar peduncles. Surprisingly, the reverse correlation between the SP Auditory measure and RD of the MCP was observed for TDC compared to the SPD children, with trends towards reversed correlations in the left and right SCP as well. More data is needed to verify these results in the TDC group that run counter to the hypothesized relationship between white matter microstructure and sensory processing. In general, the TDC group did not show significant correlations between cerebral or cerebellar peduncle WM microstructure and sensory behavior, even those with RD, FA and MD values in the same range as most SPD kids. This suggests that typically developing children are often more resilient to the potential effects of altered cerebellar white matter microstructure, perhaps because of other compensatory neural pathways. This remains an area for further exploration.

### 4.3 Cerebellar white matter microstructure and higher-order sensory behavior

Although the cerebellum has historically been implicated primarily in motor coordination, it is now clear that the cerebellum and its cortical connections are also important for higher-order processes such as multisensory integration, perceptual analysis, working memory, attention, and social cognition (Baumann et al. 2015; Van Overwalle et al. 2020; Sathyanesan, et al. 2019). As we hypothesized, significant correlations exist between white matter microstructure of the cerebellar peduncles and parent report measures of inattention and multisensory processing in the SPD cohort. The SCP, which we found to be associated with both multisensory integration and inattention SP measures, contains the cerebellothalamic tract, which transmits information via the thalamus to cerebral cortex, including higher-order association areas (Buckner 2013; Martin 2003). The MCP, which along with the SCP was associated with inattention measures in our study, receives connections from these same areas of cerebral cortex via the pons (Buckner 2013; Wang et al. 2003). Integration of auditory, somatosensory, and visual information is known to occur via mossy fibers that synapse onto cerebellar granule cells in animal experiments (Groen et al. 2011; Huang et al. 2013). While a few previous studies in humans have suggested that the cerebellum may facilitate multisensory integration (Erickson et al. 2014; Kern 2002), they have been limited to small sample sizes, including a case report of cerebellar agenesis (Ronconi et al. 2017). The current study provides additional clinical evidence that the cerebellum is involved in dysfunctional multisensory integration. Moreover, it is well established from human resting state fMRI studies that the cerebellum is an integral part of both the dorsal and ventral attention networks (Buckner et al. 2011). Our results corroborate this finding by showing altered cerebellar microstructural WM organization is related to abnormal attentional control in a clinical phenotype characterized by sensory processing dysfunction.

### 4.4 Limitations and future studies

As an initial exploration of the role of cerebellar WM microstructure in sensory processing dysfunction, this study has several limitations. To more comprehensively determine how altered WM microstructure affects sensory behavior, future studies should include a full battery of both parent report and direct observation measures. The inclusion of more objective child assessments eliminates parental bias and provides insight into which parent report measures are most reflective of directly observable sensory function. Additionally, further investigations with larger sample sizes can focus on analyzing SPD using more homogeneous subcategories based on sensory domain (e.g. tactile, auditory) and/or type of processing issue (e.g. over- or under-responsivity, seeking, discrimination). With an SPD sample as defined in the current study, it is possible to have a cohort of individuals with “opposite” sensory symptoms (e.g. sensory over-vs. under-responsivity) that might obscure important findings that could be made in each distinct subgroup. The current study was also limited in terms of certain aspects of generalizability. For example, our subjects were chosen to fall within a narrow age range prior to puberty to reduce variability in this moderate-sized cohort as well as to avoid the challenges of scanning unsedated younger children. Future longitudinal studies should include a wider age range to ensure generalizability and determine how WM integrity in SPD is affected over the course of neurodevelopment. These investigations could also use new diffusion MRI methodology such as neurite orientation dispersion and density imaging (NODDI) for more sensitive and specific evaluation of white matter microstructure (Chang et al. 2015; Owen et al. 2014; Zhang et al. 2012), including assessing children before and after various treatments, such as occupational therapy.

### 4.5 Conclusions

In summary, the current study validates previous findings of altered WM connectivity in children with SPD compared to TDC. It also demonstrates the cerebellum’s role in multisensory integration and attention in individuals with SPD. Finally, the current study contributes to the overall understanding of the neuroanatomical features present in SPD. It is important to identify these neurobiological underpinnings of the disorder, which can ultimately provide a biomarker to inform classification and diagnosis, prognostication, and interventions. It may also lend further support to SPD being considered a “brain-based” condition, rather than as a secondary symptom of ASD or a result of poor parenting, and help those with SPD obtain the knowledge and treatment options they need.

## Data Availability

Data are available for research purposes upon reasonable request to the corresponding authors.

## Conflict of Interest

The authors declare that the research was conducted in the absence of any commercial or financial relationships that could be construed as a potential conflict of interest.

## Author Contributions

Conception and design of the study: E.J.M. and P.M. Acquisition and analysis of the data: A.N., M.A.R, E.M.P., J.W-J., I.B., M.G., A.B-A. and S.D. Drafting significant portion of the manuscript and figures: A.N., M.A.R, E.M.P., E.J.M., and P.M.

## Funding

This work was funded by grants from the Wallace Research Foundation to E.J.M. and P.M., and gifts from the Mickelson-Brody Family Foundation, the Glass Family Foundation, the James Gates Family Foundation, and the Kawaja-Holcombe Family Foundation to E.J.M. Neuroimaging was supported by NIH K23 MH083890 (E.J.M.) and NIH R01 MH116950 (P.M. and E.J.M.). The Sensory, Neurodevelopment, and Autism Program (SNAP) crowdfunding contributors made this work possible.

## Acknowledgments

We profusely thank our study participants and their families, whose time and support made this work possible.

## Notes

### Competing Interest Statement

The authors have declared no competing interest.

### Author Declarations

This study was approved by the UCSF IRB and was performed in accordance with institution ethical standards.

### Summary of Updates

We updated contact info for our corresponding author

## References

Ahn, Roianne R., Lucy. J. Miller, Sharon Milberger, and Daniel N. McIntosh. 2004. “Prevalence of Parents’ Perceptions of Sensory Processing Disorders among Kindergarten Children.” The American Journal of Occupational Therapy 58 (3): 287–293. doi:10.5014/ajot.58.3.287.

Ailion, Alyssa S., Tricia Z. King, Simone R. Roberts, Brian Tang, Jessica A. Turner, Christopher M. Conway, and Bruce Crosson. 2020. “Double Dissociation of Auditory Attention Span and Visual Attention in Long-Term Survivors of Childhood Cerebellar Tumor: A Deterministic Tractography Study of the Cerebellar-Frontal and the Superior Longitudinal Fasciculus Pathways.” Journal of the International Neuropsychological Society: 1–15. doi:10.1017/S1355617720000417.

Alexander, Andrew L., Jee Eun Lee, Mariana Lazar, and Aaron S. Field. 2007. “Diffusion Tensor Imaging of the Brain.” Neurotherapeutics 4 (3): 316–329. doi:10.1016/j.nurt.2007.05.011.

American Psychiatric Association. 2013. Diagnostic and statistical manual of mental disorders: DSM-5. Arlington, VA: American Psychiatric Association.

Bar-Shalita, Tami, Jean-Jacques Vatine, Ze’ev Seltzer, and Shula Parush. 2009. “Psychophysical Correlates in Children with Sensory Modulation Disorder (SMD).” Physiology & Behavior 98 (5): 631–639. doi:10.1016/j.physbeh.2009.09.020.

Baumann, Oliver, Ronald J. Borra, James M. Bower, Kathleen E. Cullen, Christophe Habas, Richard B. Ivry, Maria Leggio, et al. 2015. “Consensus Paper: The Role of the Cerebellum in Perceptual Processes.” Cerebellum 14 (2): 197–220. doi: 10.1007/s12311-014-0627-7.

Brandes-Aitken, Anne, Joaquin A. Anguera, Camarin E. Rolle, Shivani S. Desai, Carly Demopoulos, Sasha N. Skinner, Adam Gazzaley, and Elysa J. Marco. 2018. “Characterizing Cognitive and Visuomotor Control in Children with Sensory Processing Dysfunction and Autism Spectrum Disorders.” Neuropsychology 32 (2): 148–160. doi:10.1037/neu0000404.

Brito, Adriana R., Marcio M. Vasconcelos, Romeu C. Domingues, Luiz C. Hygino da Cruz Jr, Leise de Souza Rodrigues, Emerson L. Gasparetto, and Carlos Adolfo B. Pinto Calçada. 2009. “Diffusion Tensor Imaging Findings in School-Aged Autistic Children.” Journal of Neuroimaging 19 (4): 337–343. doi:10.1111/j.1552-6569.2009.00366.x.

Bruckert, Lisa, Katherine E. Travis, Aviv A. Mezer, Michal Ben-Shachar, and Heidi M. Feldman. 2020. “Associations of Reading Efficiency with White Matter Properties of the Cerebellar Peduncles in Children.” Cerebellum. https://doi.org/10.1007/s12311-020-01162-2.

Buckner, Randy L. 2013. “The Cerebellum and Cognitive Function: 25 Years of Insight from Anatomy and Neuroimaging.” Neuron (Cambridge, Mass.) 80 (3): 807–815. doi:10.1016/j.neuron.2013.10.044.

Buckner, Randy L., Fenna M. Krienen, Angela Castellanos, Julio C. Diaz, and B. T. Thomas Yeo. 2011. “The Organization of the Human Cerebellum Estimated by Intrinsic Functional Connectivity.” Journal of Neurophysiology 106 (5): 2322–2345. doi:10.1152/jn.00339.2011.

Cardon, Garrett J., Susan Hepburn, and Donald C. Rojas. 2017. “Structural Covariance of Sensory Networks, the Cerebellum, and Amygdala in Autism Spectrum Disorder.” Frontiers in Neurology 8: 615. doi:10.3389/fneur.2017.00615.

Chang, Yi Shin, Julia P. Owen, Nicholas J. Pojman, Tony Thieu, Polina Bukshpun, Mari L. J. Wakahiro, Jeffrey I. Berman, et al. 2015. “White Matter Changes of Neurite Density and Fiber Orientation Dispersion during Human Brain Maturation.” PLOS One 10 (6): e0123656. doi:10.1371/journal.pone.0123656.

Chang, Yi-Shin, Mathilde Gratiot, Julia P. Owen, Anne Brandes-Aitken, Shivani S. Desai, Susanna S. Hill, Anne B. Arnett, Julia Harris, Elysa J. Marco, and Pratik Mukherjee. 2016. “White Matter Microstructure is Associated with Auditory and Tactile Processing in Children with and without Sensory Processing Disorder.” Frontiers in Neuroanatomy 9: 169. doi:10.3389/fnana.2015.00169/full.

Chang, Yi-Shin, Julia P. Owen, Shivani S. Desai, Susanna S. Hill, Anne B. Arnett, Julia Harris, Elysa J. Marco, and Pratik Mukherjee. 2014. “Autism and Sensory Processing Disorders: Shared White Matter Disruption in Sensory Pathways but Divergent Connectivity in Social-Emotional Pathways.” PLOS One 9 (7): e103038. doi:10.1371/journal.pone.0103038.

Dunn, Winnie. 1999. Sensory Profile: User’s Manual. San Antonio, TX: The Psychological Corporation.

Erickson, Laura C., Brandon A. Zielinski, Jennifer E. V. Zielinski, Peter E. Turkeltaub, Amber M. Leaver, and Josef P. Rauschecker. 2014. “Distinct Cortical Locations for Integration of Audiovisual Speech and the McGurk Effect.” Frontiers in Psychology. doi:10.3389/fpsyg.2014.00534/full.

Groen, Wouter B., Jan K. Buitelaar, Rutger J. van der Gaag, and Marcel P. Zwiers. 2011. “Pervasive Microstructural Abnormalities in Autism: A DTI Study.” Journal of Psychiatry and Neuroscience 36 (1): 32–40. doi:10.1503/jpn.090100.

Huang, Cheng-Chui, Ken Sugino, Yasuyuki Shima, Caiying Guo, Suxia Bai, Brett D. Mensh, Sacha B. Nelson, and Adam W. Hantman. 2013. “Convergence of Pontine and Proprioceptive Streams Onto Multimodal Cerebellar Granule Cells.” eLife 2. doi:10.7554/eLife.00400.

Jeong, Jeong-Won, Diane Chugani, Michael Behen, Vijay Tiwari, and Harry Chugani. 2012. “Altered White Matter Structure of the Dentatorubrothalamic Pathway in Children with Autistic Spectrum Disorders.” The Cerebellum 11 (4): 957–971. doi:10.1007/s12311-012-0369-3.

Kern, J. K. 2002. “The Possible Role of the Cerebellum in Autism/PDD: Disruption of a Multisensory Feedback Loop.” Medical Hypotheses 59 (3): 255–260. doi:10.1016/S0306-9877(02)00212-8.

Koziol, Leonard F., Deborah Ely Budding, and Dana Chidekel. 2011. “Sensory Integration, Sensory Processing, and Sensory Modulation Disorders: Putative Functional Neuroanatomic Underpinnings.” Cerebellum (London, England) 10 (4): 770–792. doi:10.1007/s12311-011-0288-8.

Lord, C., M. Rutter, S. Goode, J. Heemsbergen, H. Jordan, L. Mawhood, and E. Schopler. 1989. “Autism Diagnostic Observation Schedule: A Standardized Observation of Communicative and Social Behavior.” Journal of Autism and Developmental Disorders 19 (2): 185. doi:10.1007/BF02211841.

Mallott, Jacob M., Eva M. Palacios, Jun Maruta, Jamshid Ghajar, and Pratik Mukherjee. 2019. “Disrupted White Matter Microstructure of the Cerebellar Peduncles in Scholastic Athletes After Concussion.” Frontiers in Neurology 10: 518. doi:10.3389/fneur.2019.00518.

Marco, Elysa J., Leighton B. N. Hinkley, Susanna S. Hill, and Srikantan S. Nagarajan. 2011. “Sensory Processing in Autism: A Review of Neurophysiologic Findings.” Pediatric Research 69 (5 Part 2): 48R–54R. doi:10.1203/pdr.0b013e3182130c54.

Martin, John H. 2003. Neuroanatomy: Text and Atlas. 3rd ed. London: McGraw-Hill Professional.

McMahon, Kibby, Deepika Anand, Marissa Morris-Jones, and M. Zachary Rosenthal. 2019. “A Path from Childhood Sensory Processing Disorder to Anxiety Disorders: The Mediating Role of Emotion Dysregulation and Adult Sensory Processing Disorder Symptoms.” Frontiers in Integrative Neuroscience 13: 22. doi:10.3389/fnint.2019.00022.

Miller, Lucy J., Marie E. Anzalone, Shelley J. Lane, Sharon A. Cermak, and Elizabeth T. Osten. 2007. “Concept Evolution in Sensory Integration: A Proposed Nosology for Diagnosis.” The American Journal of Occupational Therapy 61 (2): 135–140. doi:10.5014/ajot.61.2.135.

Miller, Lucy J. 2009. “Perspectives on Sensory Processing Disorder: A Call for Translational Research.” Frontiers in Integrative Neuroscience 3: 22. doi:10.3389/neuro.07.022.2009.

Molholm, Sophie, Jeremy W. Murphy, Juliana Bates, Elizabeth M. Ridgway, and John J. Foxe. 2020. “Multisensory Audiovisual Processing in Children with a Sensory Processing Disorder (I): Behavioral and Electrophysiological Indices Under Speeded Response Conditions.” Frontiers in Integrative Neuroscience 14: 4. doi:10.3389/fnint.2020.00004.

Oldehinkel, Marianne, Maarten Mennes, Andre Marquand, Tony Charman, Julian Tillmann, Christine Ecker, Flavio Dell’Acqua, et al. 2019. “Altered Connectivity between Cerebellum, Visual, and Sensory-Motor Networks in Autism Spectrum Disorder: Results from the EU-AIMS Longitudinal European Autism Project.” Biological Psychiatry: Cognitive Neuroscience and Neuroimaging 4: 260–270. doi:10.1016/j.bpsc.2018.11.010.

Owen, Julia P., Yi Shin Chang, Nicholas J. Pojman, Polina Bukshpun, Mari L. J. Wakahiro, Elysa J. Marco, Jeffrey I. Berman, et al. 2014. “Aberrant White Matter Microstructure in Children with 16p11.2 Deletions.” The Journal of Neuroscience 34 (18): 6214–6223. doi:10.1523/JNEUROSCI.4495-13.2014.

Owen, Julia P., Elysa J. Marco, Shivani Desai, Emily Fourie, Julia Harris, Susanna S. Hill, Anne B. Arnett, and Pratik Mukherjee. 2013. “Abnormal White Matter Microstructure in Children with Sensory Processing Disorders.” NeuroImage Clinical 2: 844–853. doi:10.1016/j.nicl.2013.06.009.

Parsons, Lawrence M., Augusto Petacchi, Jeremy D. Schmahmann, and James M. Bower. 2009. “Pitch Discrimination in Cerebellar Patients: Evidence for a Sensory Deficit.” Brain Research 1303: 84–96. doi:10.1016/j.brainres.2009.09.052.

Payabvash, Seyedmehdi, Eva M. Palacios, Julia P. Owen, Maxwell B. Wang, Teresa Tavassoli, Molly Gerdes, Anne Brandes-Aitken, Elysa J. Marco, and Pratik Mukherjee. 2019. “Diffusion Tensor Tractography in Children with Sensory Processing Disorder: Potentials for Devising Machine Learning Classifiers.” NeuroImage Clinical 23: 101831. doi:10.1016/j.nicl.2019.101831.

Richard, Gail J. and Jeanane M. Ferre. 2006. Differential Screening Test for Processing. East Moline, IL: LinguiSystems, Inc.

Ronconi, L., L. Casartelli, S. Carna, M. Molteni, F. Arrigoni, and R. Borgatti. 2017. “When One is enough: Impaired Multisensory Integration in Cerebellar Agenesis.” Cerebral Cortex (New York, N.Y. 1991) 27 (3): 2041–2051. doi:10.1093/cercor/bhw049.

Rutter, Michael, Anthony Bailey, and Catherine Lord. 2003. The Social Communication Questionnaire: Manual. Los Angeles, CA: Western Psychological Services.

Ryckman, Justin, Claudia Hilton, Cynthia Rogers, and Roberta Pineda. 2017. “Sensory Processing Disorder in Preterm Infants during Early Childhood and Relationships to Early Neurobehavior.” Early Human Development 113: 18–22. doi:10.1016/j.earlhumdev.2017.07.012.

Sathyanesan, Aaron, Joy Zhou, Joseph Scafidi, Detlef H. Heck, Roy V. Sillitoe, and Vittorio Gallo. 2019. “Emerging Connections between Cerebellar Development, Behaviour and Complex Brain Disorders.” Nature Reviews. Neuroscience 20 (5): 298–313. doi:10.1038/s41583-019-0152-2.

Shukla, Dinesh K., Brandon Keehn, Alan J. Lincoln, and Ralph-Axel Müller. 2010. “White Matter Compromise of Callosal and Subcortical Fiber Tracts in Children with Autism Spectrum Disorder.” Journal of the American Academy of Child and Adolescent Psychiatry 49 (12): 1269–1278e2. doi:10.1097/00004583-201012000-00012.

Van Overwalle, Frank, Mario Manto, Zaira Cattaneo, Silvia Clausi, Chiara Ferrari, John D. E. Gabrieli, Xavier Guell, et al. 2020. “Consensus Paper: Cerebellum and Social Cognition.” Cerebellum. doi:10.1007/s12311-020-01155-1.

Wang, Fei, Zhiguo Sun, Xiangke Du, Xilin Wang, Zhong Cong, Hongyan Zhang, Dai Zhang, and Nan Hong. 2003. “A Diffusion Tensor Imaging Study of Middle and Superior Cerebellar Peduncle in Male Patients with Schizophrenia.” Neuroscience Letters 348 (3): 135–138. doi:10.1016/S0304-3940(03)00589-5.

Wechsler, David. 2003. Wechsler Intelligence Scale for Children–Fourth Edition (WISC-IV) Administration and Scoring Manual. San Antonio, TX: The Psychological Corporation.

Wickremasinghe, A. C., E. E. Rogers, B. C. Johnson, A. Shen, A. J. Barkovich, and E. J. Marco. 2013. “Children Born Prematurely have Atypical Sensory Profiles.” Journal of Perinatology 33 (8): 631–635. doi:10.1038/jp.2013.12.

Zhang, Hui, Torben Schneider, Claudia A. Wheeler-Kingshott, and Daniel C. Alexander. 2012. “NODDI: Practical in Vivo Neurite Orientation Dispersion and Density Imaging of the Human Brain.” NeuroImage 61 (4): 1000–1016. doi:10.1016/j.neuroimage.2012.03.072.

Zhao, Guihu, Kirwan Walsh, Jun Long, Weihua Gui, and Kristina Denisova. 2018. “Reduced Structural Complexity of the Right Cerebellar Cortex in Male Children with Autism Spectrum Disorder.” PloS One 13 (7): e0196964. doi:10.1371/journal.pone.0196964.

Zonouzi, Marzieh, Joseph Scafidi, Peijun Li, Brian McEllin, Jorge Edwards, Jeffrey L. Dupree, Lloyd Harvey, et al. 2015. “GABAergic Regulation of Cerebellar NG2 Cell Development is Altered in Perinatal White Matter Injury.” Nature Neuroscience 18 (5): 674–682. doi:10.1038/nn.3990.

